# Impact of COVID-19 related social isolation on behavioral outcomes in young adults

**DOI:** 10.1101/2022.10.20.22280791

**Authors:** Alessandra Patrono, Azzurra Invernizzi, Donatella Placidi, Giuseppa Cagna, Stefano Calza, Manuela Oppini, Elza Rechtman, Demetrios M. Papazaharias, Abraham Reichenberg, Roberto G. Lucchini, Maurizio Memo, Elisa Ongaro, Matteo Rota, Robert O. Wright, Stefano Renzetti, Megan K. Horton

## Abstract

Social isolation strongly affects our emotions, behavior and interactions. Worldwide, individuals experienced prolonged periods of isolation during the first wave of the COVID-19 pandemic when authorities imposed restrictions to reduce the spread of the virus. In this study, we investigated the effects of social isolation on emotional and behavioral outcomes in young adults from Lombardy, Italy, a global hotspot of COVID-19. We leverage baseline (pre-social isolation) and follow-up (mid-or post isolation) data collected from young adults enrolled in the ongoing, longitudinal Public Health Impact of Metals Exposure (PHIME) study. At baseline, 167 participants completed the ASEBA questionnaires (ASR/YSR) by weblink or in person; 65 completed the ASR between 12-18 weeks after the onset of restrictions. Using the sign test and multiple linear regression models, we examined differences in ASR scores between baseline and follow-up adjusting for sex, age, pre-pandemic IQ (Kaufman Brief Intelligence Tests; K-BIT 2), and time with social restrictions (weeks). Further, we examined interactions between sex and time in social isolation. Participants completed the ASR after spending an average of 14 weeks in social isolation (range 12-18 weeks). Thought Problems increased between baseline and follow-up (median difference 1.0; 1^st.^, 3^rd^ quartile: -1.0, 4.0; p=0.049). Among males, a longer time with social isolation (≥ 14 weeks) was associated with increased rule-breaking behaviors of 2.8 points. These results suggest the social isolation related to COVID-19 adversely impacted mental health. In particular, males seem to externalize their condition. These findings might help future interventions and treatment to minimize the consequences of social isolation experience in young adults.

## Introduction

The COVID-19 pandemic has led to unprecedented social distancing behavior to limit the spread of the virus (Anderson et al., 2020; Schulze et al., 2022). These measures impacted the general population, not only those found to be infected or exposed to the disease. Social distancing measures can effectively counter the spread of the disease(Glass et al., 2006), but can have an unprecedented impact on mental health and psychological well-being.

In Italy, the Lombardy area was the epicenter of the infection and one of the first places in the Western world confronted with COVID-19. No medications or vaccinations were available during the first wave of COVID-19. Therefore, the Italian government implemented a non-pharmacological measure referred to as lockdown, a forced and prolonged period of social restrictions, distancing and isolation (Pasca et al., 2021a; Shen et al., 2020) to stem the spread of infection. Lockdown social isolation refers to the “inadequate quality and quantity of social relationships with other people at the individual, group, community level and the wider social environment in which human interaction occurs” (Zavaleta et al., 2014). Italy was the first country to enter a COVID-19 related lockdown (*Gazzetta Ufficiale*, n.d.-a). The lockdown began in Northern Italy on February 23, 2020(*Gazzetta Ufficiale*, n.d.-b) and increasingly restrictive decrees followed gradually up to March 9, 2020 and the restraining measures were extended throughout Italy, and to March 11, 2020 In Italy, during this period, only essential activities and shops were accessible (i.e.,medical services, grocery stores), individuals were allowed to leave their homes only for demonstrated needs, such as for health reasons, shopping for basic needs and for work (if it was not possible to work from home)(*Gazzetta Ufficiale*, n.d.-c). Social gatherings were minimized or prohibited (Gorenko et al., 2021a). Although these restrictive measures successfully prevented more serious consequences of the COVID-19 pandemic, the social isolation may have resulted in mental health conditions (Pasca et al., 2021b). Extended social isolation conditions related to COVID-19 have been associated with short- and long-term psychosocial and mental health consequences among all ages of the population (Singh et al., 2020). The magnitude of the impact is influenced by many risk factors such as gender (Özdin & ş, 2020), age (Wilson et al., 2007), economic disadvantage (Aragona et al., 2020), and pre-existing health conditions (Conti et al., 2020). In general, sex (female), age (individuals between 18-30 years and over 60 years) and education (higher education), showed the highest levels of mental health problems following COVID-19 related social isolation(Volken et al., 2021; Xiong et al., 2020).

Although several studies have focused on mental health assessment in different subgroups of the population and especially investigated the effect of social restrictions in elderly (Gorenko et al., 2021b), there are few studies that have longitudinal data (baseline and follow-up) of the impact of COVID-19 related social isolation on the mental health of healthy young adults. In this study, we examine the impact of COVID-19 related social isolation on emotional and behavioral outcomes among healthy young adults living in Northern Italy (Province of Brescia), one of the first global hotspots of COVID-19. Using information on behavioral outcomes collected prior to and following participants experience of social isolation, we aim to quantify the impact of social isolation on young adult mental health to inform future interventions to minimize or eliminate the consequences of social isolation experience in healthy young adults.

## Materials and Methods

### Participants

Participants were part of the Public Health Impact of Metals Exposure (PHIME) study, an ongoing longitudinal cohort study of adolescents in the province of Brescia, Northern Italy. PHIME was designed to assess cognitive and behavioral function in adolescents and young adults with environmental exposure to neurotoxic metals. Participants were never to have received psychological or neuropsychological diagnosis. Other enrollment, inclusion and exclusion criteria for the PHIME study are described in detail elsewhere (Bauer et al., 2017; Chiu et al., 2017; Lucas et al., 2015; Lucchini et al., 2012). Upon enrollment, PHIME participants participated in a baseline in-person visit consisting of self-and interviewer assisted questionnaires capturing sociodemographic characteristics (i.e., sex, date of birth, residential address, parental education and occupation) and neurodevelopmental outcomes including the Kaufman Brief Intelligence Test-second edition (K-BIT 2)(Kaufman, 2004) for IQ and the Achenbach System of Empirically Based Assessment (ASEBA) Youth Self Report (YSR)(Achenbach & Rescorla, 2007a) or (ASEBA) Adult Self Report(ASR)(Achenbach & Rescorla, 2003) for behavioral and emotional regulation (Section 2.3). As part of the PHIME study, 167 participants (ages 19.3 years ± 2.3) completed the YSR or the ASR in person with a trained psychologist prior to the beginning of covid-related social isolation (on average the first visit was performed 81.8 ± 43.2 weeks prior the first day of lockdown). To assess the impact of social isolation on emotional and behavioral outcomes, we re-administered the ASR via an online platform (REDCap®, Research Electronic Data Capture, Vanderbilt University, Nashville, TN, USA) 12-18 weeks following the onset of lockdown measures (on average the subjects answered to the questionnaire after 13.4 ± 1.4 weeks). We distributed the web link to all 167 PHIME participants who completed the baseline assessment; 40% (65/167) of participants completed the online ASR. During the second time point, no information relating to the SES and the IQ was collected again, considering these variables stable at such a short time compared to the first administration.

Eligible participants received a detailed description of the study procedures before consenting to participate. The Institutional Review Boards at the Ethical Committee of Brescia, the Icahn School of Medicine at Mount Sinai, and the University of California, Santa Cruz approved all PHIME study protocols.

### Aseba Young and Adult Self Report (YSR and ASR) questionnaires

The Achenbach System of Empirically Based Assessment (ASEBA) offers a comprehensive approach to assessing adaptive and maladaptive functioning. During the baseline PHIME visit, adolescent participants (ages 15-17 years; n = 45) completed the Youth Self Report (YSR) questionnaire (Achenbach & Rescorla, 2007b, 2001) and adult participants (ages 18-25years; n = 122) completed the Adult Self Report (ASR) questionnaire (de Vries et al., 2020). The YSR questionnaire is designed for self-reporting in the 11-17 years age range; the ASR is appropriate for adults ages 18-59 years. ASR questionnaires evaluate the following clinical areas: I) Anxious/Depressed; II) Withdrawn; III) Somatic Complaints; IV) Thought Problems; V) Attention Problems; VI) Aggressive Behavior; VII) Rule-Breaking Behavior; VIII) Intrusive. An Internalization Problem Composite Scale is aggregated from the individual symptoms scales: anxiety (18 items), withdrawn (9 items) and somatic complaints (12 items). An Externalizing Problem Composite Scale is composed of: aggressive behavior (15 items), rule-breaking behavior (14 items) and intrusive behavior (6 items). The other scales concern attention problems (15 items) and Thought Problems (10 items). The scale Other problems (21 elements) includes elements that do not frame any syndrome. The remaining 11 items measure adaptive functioning.

The clinical scales investigated by the ASR are comparable to those of the YRS with some changes related to the adaptation of the items by age. In the YSR version the component of depression is investigated both by scale I and II; the ASR scale VIII) Intrusive, corresponds to the YSR scale V) Problems of thought.

Since the seven ASR syndromes have YSR-rated counterparts, questionnaire scores can be directly compared (Achenbach, 2019).A score from 0 (behavior / problem absent) to 2 (behavior / problem present) is applied to each item that makes up the individual scales considered in the YSR / ASR questionnaires. Each scale will then assume a numerical value which is transformed into a T score in a range from 50 to 100. Scores between 50 and 64 are considered normal; scores between 65 and 70 are considered borderline; scores above 70 are considered clinically significant.

Both the YSR and ASR yield a Total Problems score indicating the overall psychopathological assessment of the individual (a higher score indicates greater psychopathology). Symptomatic scales correlate with the DSM-oriented diagnosis (i.e., ASR depressive symptoms correlate with DSM-diagnosed depression).

#### Covariate data

Sociodemographic data (i.e., participant sex and age, and parental occupation and education) was collected at the baseline assessment through questionnaires. Intelligence quotient (IQ) was measured using the Kaufman Brief Intelligence Test, 2^nd^ edition (KBIT-2)(Kaufman & Kaufman, 2004), a short measure of verbal and non-verbal intelligence for children, adolescents and adults, aged 4 to 90 years. The scores of the verbal and non-verbal yield a composite IQ score that can be considered as a measure of general intelligence with good correlations with other tests of intellectual functioning. An index of family socioeconomic status (SES; low, medium, or high) was calculated from parental age, occupation and education(Cesana et al., 1995).

### Statistical analysis

Descriptive statistics were used to assess the distribution of variables; continuous variables are analyzed using the median and the interquartile (IQR) range (i.e. the first and the third quartiles) and for categorical variables, we used absolute frequencies and percentages. Student *t*-tests with Welch’s correction for continuous variables and Chi-squared (*χ*_*2*_) tests for categorical variables were used to examine differences in demographic characteristics across the participants. The time spent in social isolation was calculated in weeks starting from the start of social isolation (March 9, 2020) and the date the participants completed the follow-up ASR online. Time spent in isolation was then classified as low if the elapsed time was less than 14 weeks between the initiation of the first restrictions and the follow-up questionnaire response or high if the elapsed time was greater than or equal to 14 weeks between onset of social isolation and follow-up questionnaire response (Figure 1).

**Figure 1.**
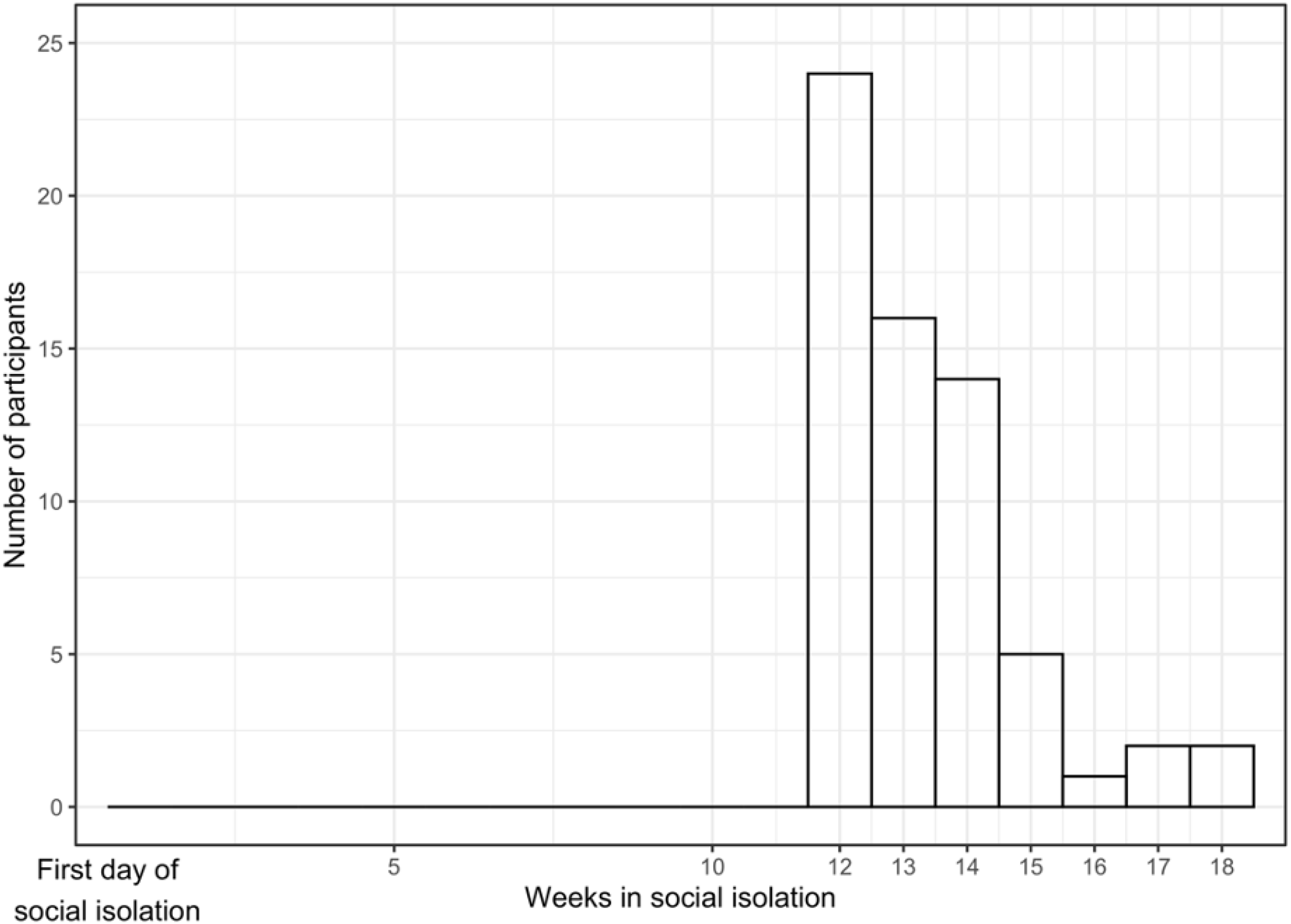
Time elapsed in social isolation. The amount of time spent in isolation was calculated as the number of weeks elapsed between the start of lockdown (3/09/2020) and the administration of the follow-up ASR and categorized as low (<14 weeks in social isolation) and high (≧14 weeks in social isolation) based on the median weeks spent in social isolation prior to assessment.

We used the sign test, a nonparametric test to assess differences between paired observations, to assess the difference between YSR/ASR symptoms before and after social isolation. The choice of the sign test was also driven by the non-symmetric distribution of differences. We then examined how the amount of time in social isolation, defined as the time elapsed between the first day of social restrictions and the follow-up visit, impact differences in YSR/ASR scores. We applied a linear regression model to examine how time elapsed in isolation (independent variable) predicted the change on YSR/ASR scores, adjusting for age, sex, baseline SES and IQ. We then determined whether the associations between time in social isolation and the change in ASR scores differed by sex through a multiplicative interaction term. Statistical significance level was set at 5% for all tests. All the statistical analyses were performed with R (version 4.1.0).

## Results

### Sociodemographic characteristics

Sociodemographic characteristics of PHIME participants included in this study are presented in Table 1. 167 participants (81 male, 19.3 ± 2.3 years) completed the initial YSR/ASR questionnaire (i.e., baseline) and 65 (39%) of these participants (26 male, 19.8 +/-2.4 years) repeated the ASR during or following the lockdown (i.e., follow up). Participants experienced an average of 14.6 +/-9.5 weeks with social restrictions before completing the follow up ASR questionnaire (Figure 1). The average IQ was 106.1 (SD 9.7); IQ did not differ between those who participated in follow up and those who did not. No YSR/ASR scores indicated problematic behaviors at baseline or follow up (Supplement table). Sociodemographic characteristics and baseline ASR scores of those participants who completed the online follow-up assessment did not differ from those who did not complete the assessment.

**Table 1.**
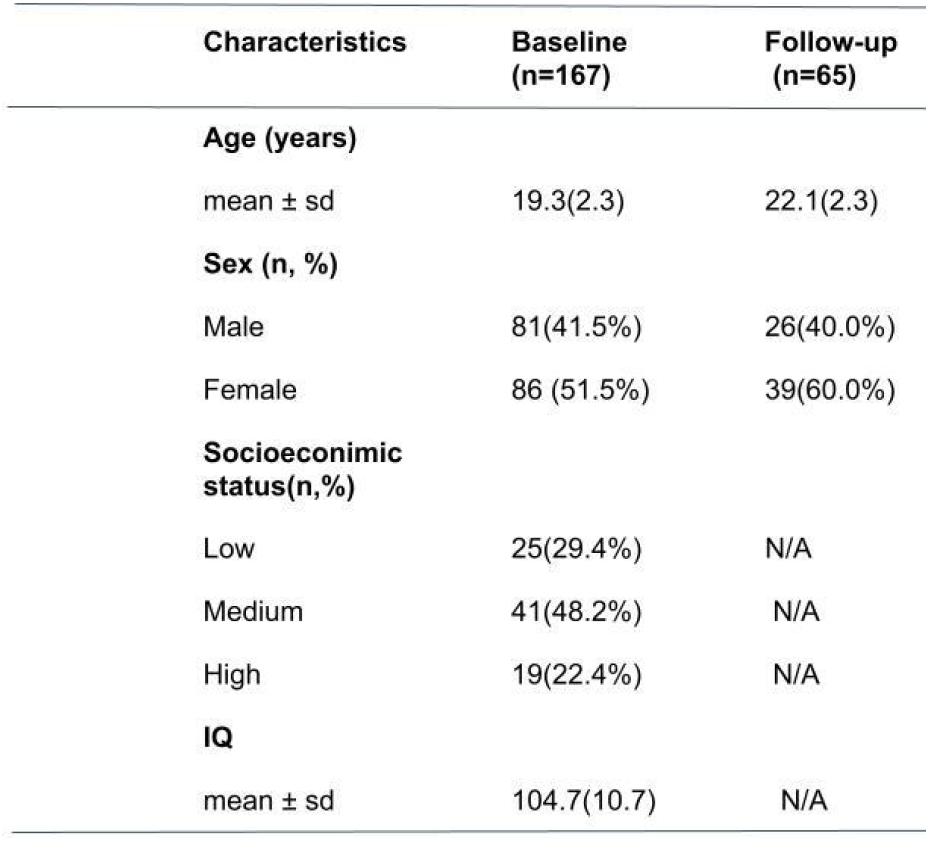
Sociodemographic characteristics of PHIME participants included in this study at baseline (n=167) and follow-up (n=65). **Note:**Mean, standard deviation (sd), range (minimum and maximum values), and percentage (%) are reported. P-values quantify differences between groups were derived using Student t-tests for continuous variables and *Χ*_*2*_ tests for categorical variables. \**p*<0.005.

### Social isolation and behavioral outcomes

We observed no differences in Total Problems reported at baseline and follow-up (Figure 2A). Participants reported significantly more Thought Problems at follow up (Sign test;51.5 (50.0, 55.8) vs 53.5. (51.0, 58.0), p = 0.049; Figure 2B). None of the other ASR scales differed significantly between baseline and follow-up (Table S1).

**Figure 2.**
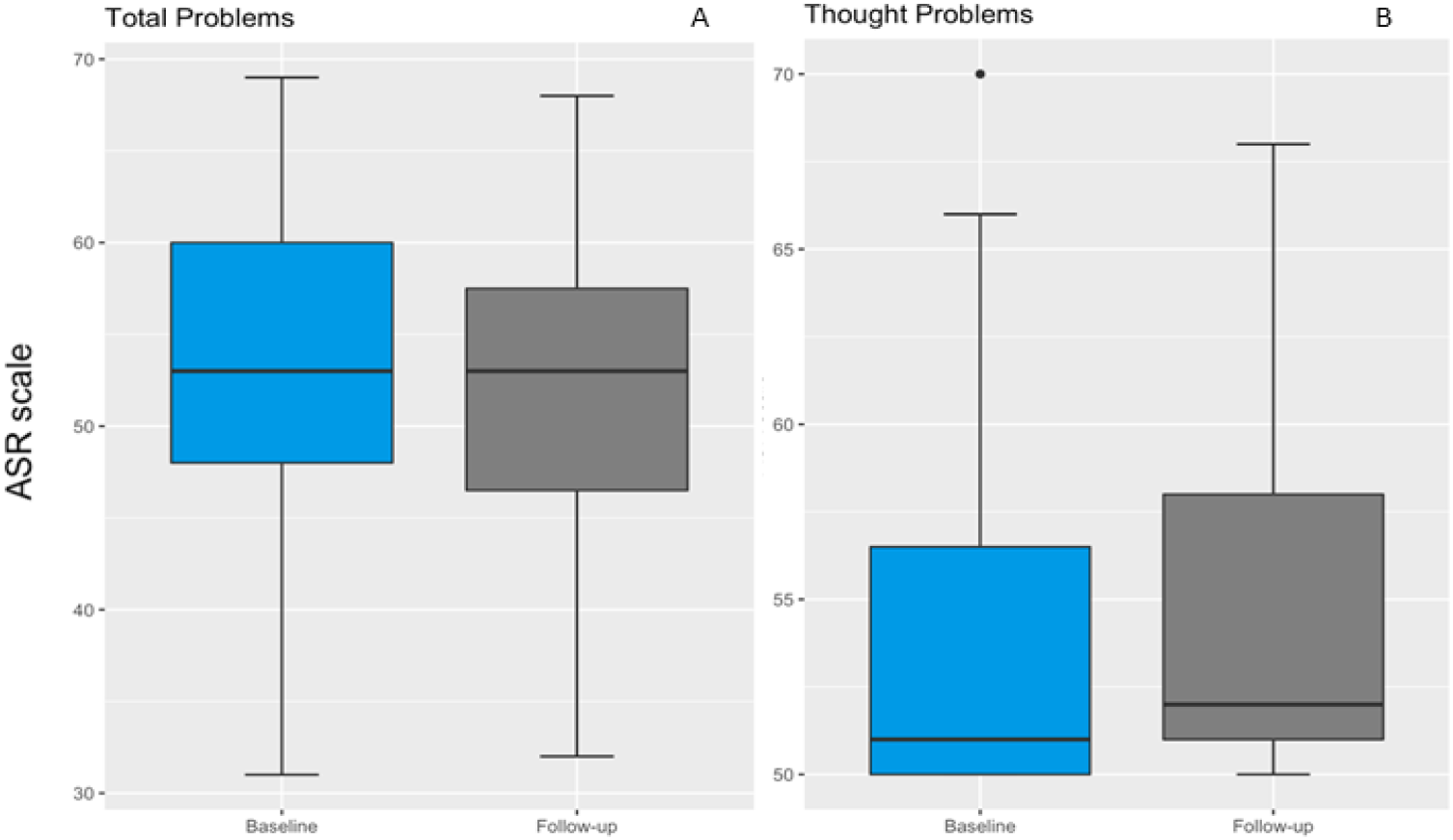
Differences in ASR score between baseline and follow up. The boxplot shows YSR/ASR scores for Total Problems (A) and Thought Problems (B) at baseline (blue box) and follow up (gray box). The error bars are the 95% confidence interval, the bottom and top of the box are the 25th and 75th percentiles, the line inside the box is the 50th percentile (median), and any outliers are shown as open circles. No differences are shown in Total Problem score scales between baseline and follow-up. Thought Problems are significantly higher in the follow-up assessment, with an overall clinical worsening in the post-social isolation period. The sign test was applied to test the difference between baseline and follow-up.

### Length of social isolation and behavioral outcomes

Spending a longer amount of time in social isolation (< 14 compared to ≧14 weeks) was marginally associated with an increase of 1.73 points in rule-breaking behaviors (linear regression, *β* 1.73; 95% Confidence Interval (CI) -0.03, -3.48 p = 0.053), after adjusting for sex, age, SES and IQ.

### Sex-specific effects of social isolation on behavioral outcomes

In the interaction between time and sex analysis, the amount of time spent in social isolation was significantly associated with increased rule-breaking behavior in males only (β = 2.8, 95%CI 0.06, 5.5; Figure 3). Male participants who spent more time in social isolation (≥ 14 weeks) reported a 3-point increase in Rule Breaking Behavior compared to males who spent less time (< 14 weeks) in social isolation. No differences in the association between the time elapsed in social isolation and ASR scores were found for female participants.

**Figure 3.**
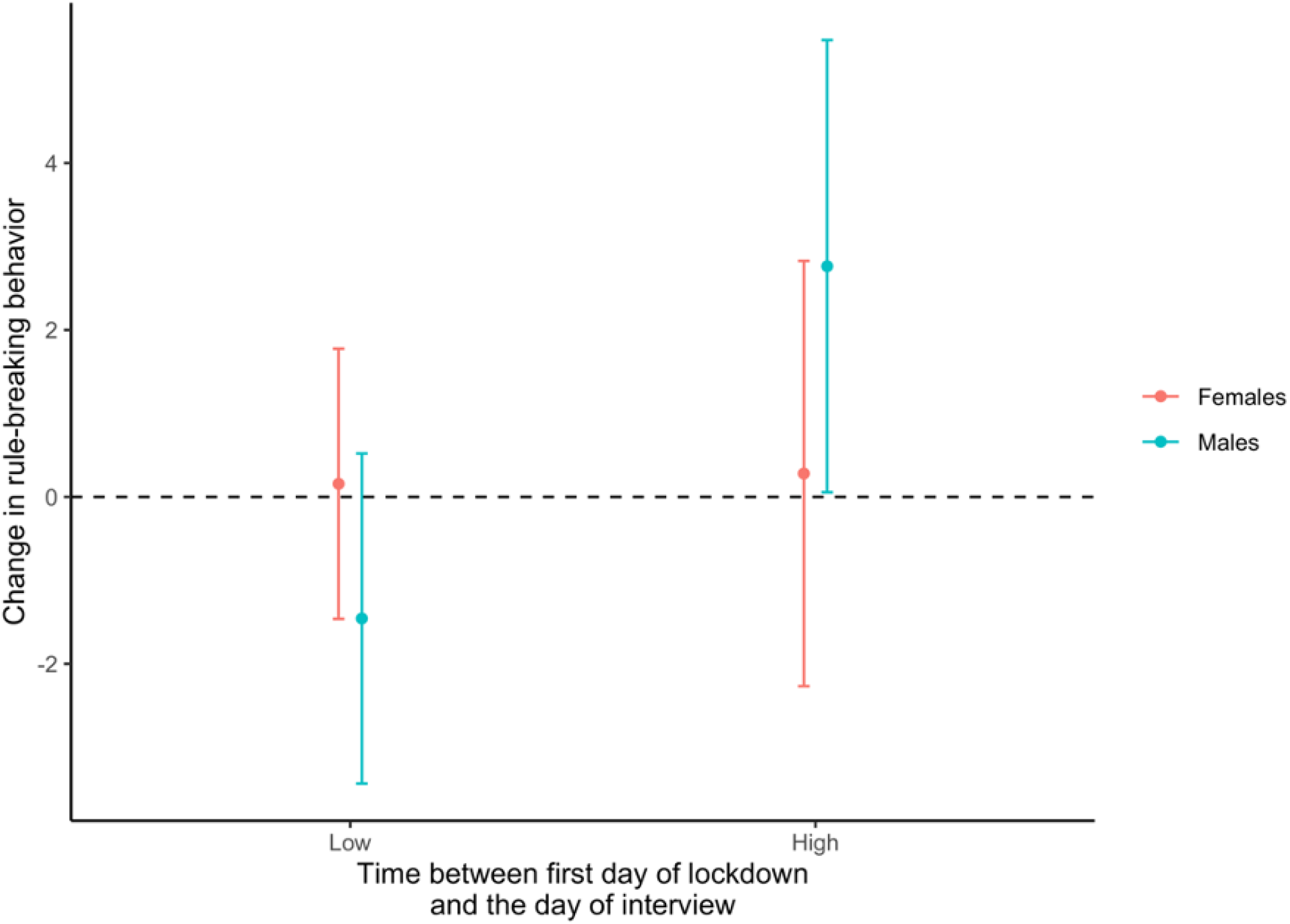
Social isolation and Rule Breaking Behavior. Results from the linear regression model including an interaction term between time in social isolation and sex. Here are displayed the marginal effects of the time spent in social isolation (< 14 weeks vs. ≧14 weeks vs low) on the difference in the ASR rule-breaking behavior score by sex. The model was adjusted by age (years), SES and IQ. The statistical significance for males with a higher time spent in social isolation is p=0.046.

## Discussion

In this study, we assess the impact of COVID-19 related social isolation on baseline (pre-isolation) and follow-up (mid-or post-isolation) behavioral outcomes in young adults enrolled in an ongoing longitudinal PHIME cohort in Northern Italy. Our findings suggest that social isolation is associated with increased Thought Problems. Further, the length of time spent in social isolation adversely impacts males compared to females; males who spend more time in social isolation reported higher Rule Breaking behavior. In our study, males reported higher Rule Breaking Behavior scores after social isolation than females. Further, spending a longer amount of time spent in social isolation increased the severity of Rule Breaking Behavior scores (i.e., more time, more rule breaking). The construct of Rule Breaking Behavior is defined as “non-compliance with the applicable regulatory expectations of the group” (Kaplan, 1980) and is related to disinhibition (Burt et al., 2008). The general construct of Rule Breaking is considered a transitory factor within the behavior and mediated by the environmental situation (Eley et al., 2003; Tackett et al., 2005). Our findings in males contribute to the literature on social isolation and behavioral outcomes as most of the previous studies focus on female mood disorders related to the pandemic and social isolation (Almeida et al., 2020; Wang et al., 2020). Our data are fairly consistent with studies that broadly analyze gender differences in typical traits in mental disorders, with a higher frequency of behavioral outcomes in males (Paris, 2007) although these series may have been influenced by bias (Braamhorst et al., 2015; Crosby & Sprock, 2004).

Our unique study design and population, located in one of the first global hotspots of the first wave of the COVID-19 pandemic, provided the opportunity to examine the impact of the pandemic related social isolation on healthy young adults. Our findings suggesting that males may be more vulnerable to the impacts of social isolation on rule breaking behavior could help direct targeted interventions. In general, males are less likely to seek therapeutic interventions to treat mental health or mood-related disorders(Liddon et al., 2018). Based on the externalizing symptomatology that drives male behavior, initiatives that focus more on attention to functioning than to emotionality should be considered. General practitioners must be instructed to differentiate gender-specific alarm bells for subsequent referral to specialist treatment.

### Limitations

The sample is relatively small and the participation rate was modest (39.88%). The low compliance could have driven a selection bias, with a possible tendency to respond to the questionnaire only by the most emotionally affected subjects. However, although the sample is not very large, it assumes importance due to the possibility of being able to compare the scores with the previous administration of the questionnaire. Another limitation is the lack of information on COVID-19 infection and its possible impact of emotional and behavioral outcomes. At the time of our follow-up, accessibility to antigen and antibody verification of the presence of the disease was limited and took place only in the presence of symptoms. Rapid testing was not widespread.

## Conclusion

To conclude, this study demonstrates how COVID-19 social restrictions policies negatively impacted on mental and behavioral health in healthy young adults. The worsening of clinical scales of ASR, with regard to the pandemic period, is theoretically and clinically significant and reflects the need to implement intervention dynamics aimed at containing or preventing long-term effects of social isolation. Future studies are needed to understand how targeted interventions, based on the results of this and other similar research pieces, can address changes in public health wellbeing.

## Data Availability

All data produced in the present work are contained in the manuscript

## Acknowledgements

The authors would like to acknowledge support from the National Institutes of Environmental Health Sciences (NIEHS) grants numbers R01 ES019222, R01 ES013744, P30ES023515, and the European Union through its Sixth Framework Programme for RTD (contract number FOOD-CT-2006-016253).

## Supplementary Materials

**Table S1.** Complete ASR scores at the baseline and follow-up visit. ASR scores (median, 1^st^ and 3^rd^ quartile) obtained baseline and follow-up and the differences estimated between the two time-points for 65 PHIME participants. P-values are estimated through the sign test

| **ASR score** | **Baseline** | **Follow-up** | **ASR pre-post** | **p** |
| --- | --- | --- | --- | --- |
| Anxious Depressed | 58.0 (53.0, 65.0) | 58.0 (52.0, 63.2) | 0.0 (-3.2, 3.0) | 1.000 |
| Withdrawn | 55.0 (51.0, 65.0) | 55.0 (50.0, 63.0) | -1.0 (-4.0, 3.0) | 0.108 |
| Somatic Complaints | 56.0 (51.0, 62.0) | 54.0 (51.0, 58.0) | 0.0 (-4.0, 2.0) | 0.480 |
| Thought Problems | 51.5 (50.0, 55.8) | 53.5 (51.0, 58.2) | 1.0 (-1.0, 4.0) | 0.049 |
| Attention Problems | 54.0 (51.0, 58.0) | 55.0 (51.0, 58.2) | 0.0 (-2.0, 3.0) | 1.000 |
| Rule Breaking Behavior | 51.0 (50.0, 53.0) | 51.0 (50.0, 52.0) | 0.0 (-1.0, 0.0) | 0.188 |
| Aggressive Behavior | 52.5 (50.0, 57.0) | 53.0 (51.0, 58.0) | 0.0 (-2.0, 3.0) | 0.784 |
| Internalizing Problems | 59.0 (52.0, 66.0) | 58.0 (51.0, 64.0) | -2.0 (-6.2, 4.0) | 0.124 |
| Externalizing Problems | 51.5 (42.8, 55.0) | 50.0 (43.8, 54.2) | 0.0 (-5.0, 4.0) | 1.000 |
| Total Problems | 52.5 (48.0, 60.0) | 53.0 (46.8, 58.0) | 0.0 (-4.2, 3.0) | 0.590 |

**Figure S1.**
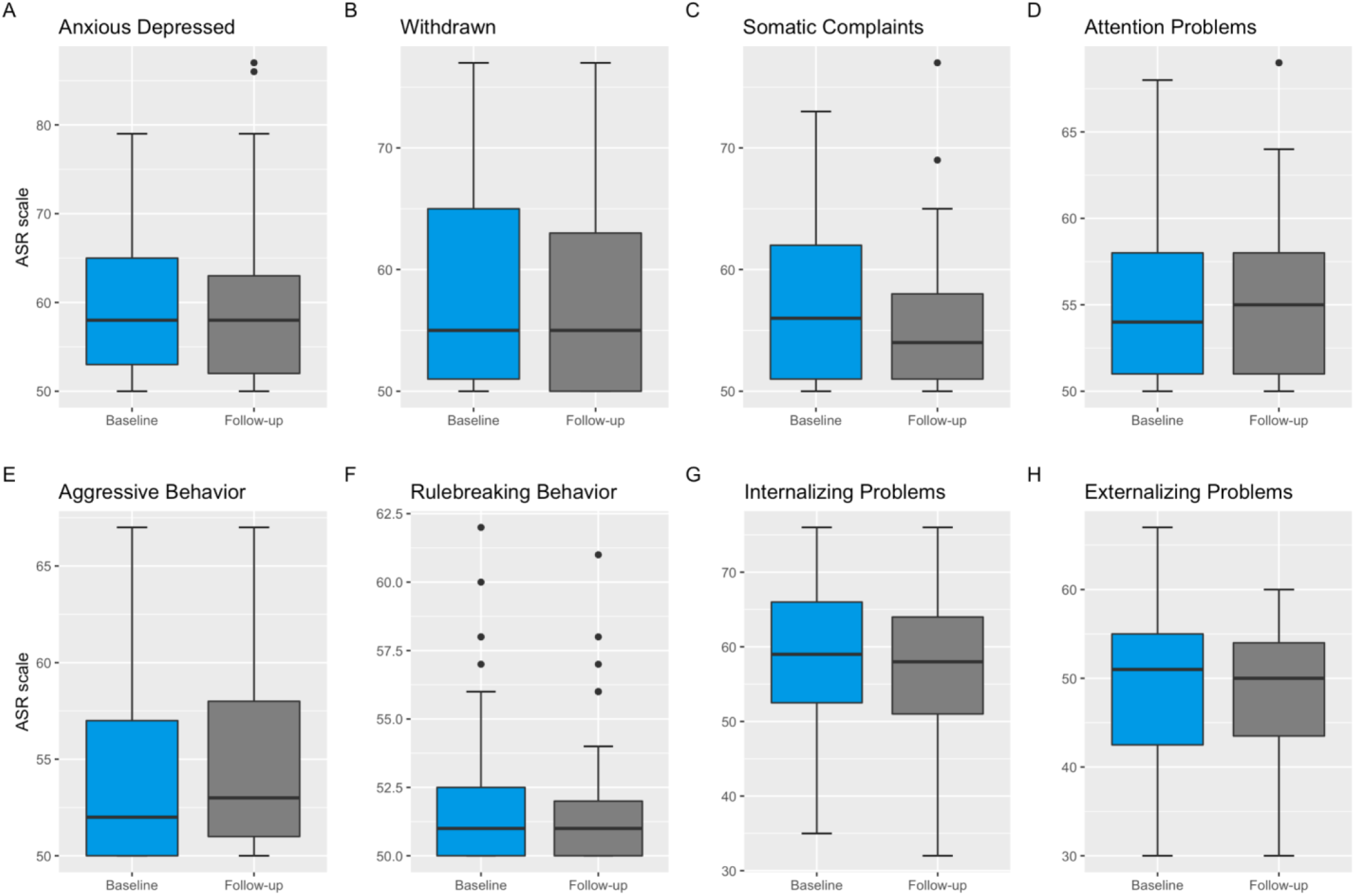
Differences in all ASR score between baseline and follow up. The boxplot shows the scores obtained in six clinical scales measured by the ASR and in “Internalizing “ and “Externalizing” problems scale of the questionnaire in the two time points considered: baseline visit (blue box) and follow up visit (gray box).

**Table S2.** Interaction between socio-demographic variables and ASR scales. Relationship between the sociodemographic variables (sex age, SES and IQ) of the sample and the psychopathological outcomes at the follow-up visit.

| *Predictors* | **Anxious Depressed** |  |  | **Withdrawn** |  |  | **Somatic Complaints** |  |  | **Thought Problems** |  |  | **Attention Problems** |  |  |
| --- | --- | --- | --- | --- | --- | --- | --- | --- | --- | --- | --- | --- | --- | --- | --- |
|  | *Estimates* | *CI* | *p* | *Estimates* | *CI* | *p* | *Estimates* | *CI* | *p* | *Estimates* | *CI* | *p* | *Estimates* | *CI* | *p* |
| High vs Low SI | 0.62 | -2.91 – 4.16 | 0.725 | -0.32 | -4.43 – 3.79 | 0.877 | -1.27 | -4.51 – 1.98 | 0.438 | 0.1 | -2.66 – 2.86 | 0.943 | -0.22 | -2.85 – 2.40 | 0.866 |
| Sex: Females vs Males | 0.83 | -2.34 – 4.00 | 0.602 | -1.04 | -4.58 – 2.51 | 0.559 | 2.8 | -0.11 – 5.72 | 0.059 | -0.23 | -2.71 – 2.24 | 0.852 | 1.38 | -0.98 – 3.73 | 0.247 |
| SES: Medium vs Low | 1.92 | -2.06 – 5.90 | 0.338 | -0.1 | -4.73 – 4.52 | 0.964 | -0.71 | -4.36 – 2.94 | 0.700 | -0.47 | -3.58 – 2.63 | 0.761 | -0.75 | -3.70 – 2.20 | 0.613 |
| SES: High vs Low | 1.21 | -3.38 – 5.80 | 0.600 | -4.21 | -9.55 – 1.14 | 0.120 | -1.29 | -5.50 – 2.92 | 0.542 | -2.18 | -5.76 – 1.40 | 0.228 | 0 | -3.41 – 3.41 | 0.999 |
| Age | -0.08 | -0.75 – 0.59 | 0.812 | 0.11 | -0.82 – 1.05 | 0.809 | 0.59 | -0.02 – 1.21 | 0.060 | 0.29 | -0.23 – 0.82 | 0.270 | 0.31 | -0.19 – 0.81 | 0.214 |
| IQ | 0.17 | 0.01 – 0.34 | **0.04** | 0.22 | 0.05 – 0.39 | **0.012** | 0.06 | -0.09 – 0.21 | 0.460 | 0.05 | -0.07 – 0.18 | 0.413 | 0.13 | 0.00 – 0.25 | **0.043** |
| *Predictors* | **Rulebreaking Behavior** |  |  | **Aggressive Behavior** |  |  | **Internalizing Problems** |  |  | **Externalizing Problems** |  |  | **Total Problems** |  |  |
|  | *Estimates* | *CI* | *p* | *Estimates* | *CI* | *p* | *Estimates* | *CI* | *p* | *Estimates* | *CI* | *p* | *Estimates* | *CI* | *p* |
| High vs Low SI | 1.73 | -0.03 – 3.48 | 0.053 | 1.21 | -1.54 – 3.95 | 0.381 | -0.77 | -4.73 – 3.19 | 0.700 | 2.2 | -2.07 – 6.47 | 0.306 | 0.6 | -2.67 – 3.87 | 0.714 |
| Sex: Females vs Males | 0.48 | -1.09 – 2.05 | 0.542 | 0.06 | -2.40 – 2.53 | 0.959 | 0.82 | -2.74 – 4.37 | 0.647 | 0.25 | -3.58 – 4.07 | 0.898 | 0.49 | -2.44 – 3.42 | 0.740 |
| SES: Medium vs Low | -1.14 | -3.11 – 0.83 | 0.249 | -0.11 | -3.20 – 2.98 | 0.943 | 2.2 | -2.25 – 6.65 | 0.327 | 2.7 | -2.10 – 7.49 | 0.265 | 1 | -2.67 – 4.67 | 0.588 |
| SES: High vs Low | -1.36 | -3.63 – 0.92 | 0.237 | 0.32 | -3.25 – 3.88 | 0.860 | -0.88 | -6.01 – 4.26 | 0.733 | 2.6 | -2.94 – 8.14 | 0.351 | -0.9 | -5.14 – 3.34 | 0.672 |
| Age | 0.35 | 0.02 – 0.68 | **0.04** | -0.13 | -0.66 – 0.39 | 0.611 | -0.07 | -0.82 – 0.69 | 0.859 | -0.19 | -1.01 – 0.62 | 0.634 | 0.2 | -0.42 – 0.83 | 0.514 |
| IQ | 0.04 | -0.04 – 0.12 | 0.342 | 0.12 | -0.00 – 0.25 | 0.055 | 0.18 | -0.01 – 0.36 | 0.060 | 0.13 | -0.07 – 0.33 | 0.189 | 0.15 | 0.00 – 0.30 | **0.048** |

